# The contribution of case mix, skill mix and care processes to the outcomes of community hospitals: a population-based observational study

**DOI:** 10.1101/2020.06.27.20141010

**Authors:** Davide Pianori, Kadjo Yves Cedric Adja, Jacopo Lenzi, Giulia Pieri, Andrea Rossi, Maria Pia Fantini

## Abstract

**Background:** New organizational models to face the unmet needs of frail patients are needed. Community hospitals (CHs) could foster integration between acute and primary care. The aim of this study was to investigate which patients’ characteristics and which care processes affect clinical outcomes, in order to identify who could benefit the most from CH care.

**Methods:** This study included all patients aged ≥65 and discharged in 2017 from the 16 CHs of Emilia-Romagna, Italy. Data sources were the regional CH informative system and hospital discharge records. CH skill mix and processes of care were collected with a survey. The study outcome was variation of the Barthel index (BI). We performed a 2-level random-intercept logistic regression analysis, and used the variance partition coefficient (VPC) to quantify the proportion of BI improvement that lay at CH level.

**Results:** Of the 13 CHs, 8 had a general practitioner medical support model, and 6 had >12 nurses’ working hours/week/bed. Overall, 53% of the patients had a BI improvement ≥10. The patient case mix explained a portion of variability across CHs. Skill mix and processes of care were not associated with BI change.

**Conclusions:** Patients’ characteristics explained part of between-CH variation in BI improvement. Professional skill mix and processes of care, albeit fundamental to achieve appropriate care and respond to the unmet needs of the frail elderly, did not account for differences in CH-specific outcomes.

## INTRODUCTION

Due to longer life expectancy and lower birth rates, currently all Western countries are facing the aging of populations and the increase of old-age dependency ratio. The EU predicts that by 2060, between 22% and 36% of all citizens will be ≥65 years old, with 12% of citizens being ≥80 years [1]. The increase in longevity in all countries of the world [2] has amplified both the prevalence of patients with two or more chronic diseases (multimorbidity) and the request for complex long-term care for emerging health-care needs [3 - 7]. Older adults are increasingly facing disability, multimorbidity and chronic illnesses, such as diabetes, hypertension, and dementia [8], which indicates a shift from acute to chronic diseases [9]. Hence, the number of home-dwelling individuals depending on healthcare and social services will continue to increase.

Most health systems are not yet sufficiently equipped with organizational tools and models to face the challenges raised by these new care needs [10], and a range of services and models are being developed worldwide to find a solution for delivering care closer to home [11, 15].

Intermediate care has been developed since the late 1990s, to foster the integration between acute and primary care in several European countries, as a broad range of services, particularly for elderly and frail patients who have complex support needs and circumstances, with the objective to deliver care meeting those needs and to achieve greater system efficiency [16].

Intermediate care has both “preventive” aims (avoiding unnecessary hospitalizations and “delayed discharges”) and “rehabilitation” aims (supporting discharge and access to rehabilitation services to enforce care close to home) [17, 18].

Community hospitals (CHs) are a bed-based intermediate-care service defined as local hospitals staffed mainly by general practitioners (GPs) and nurses to provide care in a hospital setting. The notion of a CH has evolved over time, with a diversity of service-delivery models developing in response to the needs of the local populations and in the context of a broader change in the nature of the delivery of health-care services themselves [19].

In Italy, the Ministerial Decree № 70/2015 defined CHs as facilities with a limited number of beds (≤20) managed by nurses and general practitioners or specialists intended for patients with stable clinical conditions, needing care that cannot be provided at home, but not needing the intensity of care of acute hospitals. Patients can be admitted from hospital wards, emergency rooms, homes, or residential care facilities for the elderly [20]. The recommended length of stay in the facilities is three weeks, extendible to six for cases who need particular care. CHs’ beds can be located in acute hospitals, residential facilities or health-care homes (*case della salute*), i.e. community-based facilities which offer health and social services to the population residing in the area. Health-care homes are managed by the local primary care department and rely on strong collaboration among different professionals to guarantee continuous and holistic care.

Italian regions must provide the so called “essential levels of care” (i.e., the core benefit package and standard of health services), set by the central government, through the regional health services, but they are responsible for the local organization and delivery of health care [21, 22]. Emilia-Romagna (northern Italy) still lacks tailored CH regional guidelines; however, on June 1, 2020, 23 CHs were in operation, a higher number of those present at the moment of our study.

Given the diversity of clinical conditions (case mix) of the patients who can be admitted to the CHs, the professional figures (skill mix) involved in these settings are likely to vary based on the population’s needs, context and purpose of the CH [23]. The debate is ongoing on who are the patients most suited to receive care in the CH and consequently who are the professionals most appropriate to obtain the best patient’s outcomes [24]. To date studies have shown mixed results on the impact of intermediate care services and their organization on patients’ outcomes, but there is growing evidence that these services can improve function in the elderly [25].

The aim of this study was to assess to which extent patients’ clinical conditions and care processes affect health outcomes, in order to identify who can benefit the most from receiving care in CHs and the best skill-mix to deliver care in this particular setting.

## MATERIALS AND METHODS

### Study population

This retrospective observational study included all patients residing in Emilia-Romagna discharged in 2017 from the 16 CHs that were in operation at that time in the region. In case of multiple discharges for the same patient, the first episode was retained in the analyses. Data were retrieved from the Regional Informative System of Community Hospitals (*Sistema Informativo Regionale Ospedali di Comunità, SIRCO*), which includes demographics, source of admission, cause of hospitalization, Barthel score on admission and discharge, length of stay, and destination at discharge. In the informative system, each patient is assigned a pseudonymized identification number, which remains the same across all health-care administrative databases of Emilia-Romagna.

To provide an overview of patients’ medical histories, additional clinical information was collected from primary and secondary diagnoses of all acute hospital discharges occurring 3-years prior to the CH admission (source: hospital discharge records). Specifically, we identified 31 conditions based on the Elixhauser method [26], plus myocardial infarction (ICD-9-CM codes 410.x, 412), cerebrovascular diseases (362.34, 430.x–438.x), dementia (290.x, 293.x, 294.x, 310.x,, 331.2), and leukemia (204.x-208.x). We also detected the presence of hip fractures occurring 30-day prior to the index admission (820.x-829.x).

### Characteristics of community hospitals

Data on skill mix and processes of care were collected using an ad hoc survey. The questionnaire contained 34 questions divided into 4 categories (general information, clinical activities, nursing activities, rehabilitation activities). Questions were either multiple-choice or yes–no questions. The general information section investigated responsible authority, number of beds and location (health-care home vs. acute hospital). The clinical activities section investigated medical support (GP vs. specialist), work shifts (i.e., doctors involved on weekdays, weekends, off days and nights) and availability of specialists’ consultations. The nursing activity section investigated the presence of case managers (i.e., nurses who manage the entire clinical pathway of the patients), work shifts for nurses (number of members per shift and weekly working hours). The rehabilitation activity section investigated the presence of physiotherapists, other health professionals and gyms.

The questionnaires were sent by email in May 2018 to the chief medical officers of the local health-care authorities and to the hospital trust responsible for the CH and were collected through August 2018. Thirteen out of the sixteen community hospitals operating at the time in the region responded to the questionnaire.

### Outcome measures

The study outcomes were:

- In-hospital variation of the Barthel index (≥10 vs. <10 points)
- Destination at discharge (acute hospital vs. home or residential care facility)
- CH length of stay (<20 vs. ≥20 days).

The Barthel index is a scale that measures a patient’s degree of independence; its score ranges from 0 to 100 and is inversely proportional to the patient’s degree of disability. Continuous measures were transformed into binary data to simplify the analysis and interpretation of the results.

### Exclusion criteria

Because the clinical presentation and progress of younger patients can differ from those of patients who are typically referred to intermediate care services, subjects aged <65 years were excluded from the analyses. Other exclusion criteria were:

- Barthel index score on admission >90
- Death during CH stay.

### Statistical analysis

A set of multivariable analyses was carried out to assess the association of the study outcomes with the case mix, skill mix and processes of care of CHs.

To estimate the impact of patient- and hospital-level predictors and to account for clustering, a two-level random-intercept logistic regression analysis was performed. Due to the limited number of clusters (*n* = 13), these models were estimated using the Bayesian Markov chain Monte Carlo (MCMC) instead of the default frequentist approach [27]. Briefly, Bayesian methods do not rely on asymptotic sample sizes to produce unbiased estimates, and treat parameters as random variables rather than fixed quantities. As a result, parameters have posterior distributions rather than a single point estimate, which depend on prior distributions that have to be specified before the model is run.

In our models, we used zero-mean normal priors for regression coefficients and inverse-gamma prior for the variance of random intercept (i.e., variability across providers). Because prior distributions have an increased effect on posterior distributions when sample sizes are smaller, we attempted an alternate model estimation using a half-Cauchy prior for the variance component [28], but results did not change appreciably (data not shown). In each regression model we specified 20,000 MCMC samples with a burn-in period of 5,000 to ensure convergence for all model parameters, and set a thinning interval of 5 to decrease the autocorrelation of the simulated samples. Posterior distributions of regression coefficients, including the shrinkage estimators of hospital-specific random effects, were summarized with exponentiated means (posterior odds ratios [ORs]) and with 2.5th and 97.5th percentiles (95% credible intervals). Interpretation of these quantities does not differ much from interpretation of their frequentist counterparts. The posterior distribution of random-intercept variance was summarized via the variance partition coefficient (VPC), which quantifies the proportion of total outcome variance which lies at the hospital level. The VPC ranges from 0 to 1, and a higher VPC denotes a higher variability in outcomes across hospitals [29].

To determine the portion of variability across hospitals accounted for by patient case mix and organizational variables, three distinct Bayesian multilevel models were built for each study outcome. The first model had no explanatory variables, the second model included patient-level covariates (case mix), and the third model included both patient- and hospital-level covariates (case mix, skill mix and process of care). If the VPC decreases when covariates are added to the model, part of the variation across hospitals is accounted for by these variables. To aid VPC interpretation, estimates of hospital-specific odds ratios and their 95% credible intervals resulting from multilevel regression models were plotted around the null value of 1, which indicates no difference relative to the overall weighted odds. These estimates arise from the shrinkage estimates of hospital-specific random effects, and are similar to the ratio of observed-to-expected rates (O/E), which represents the hospital risk relative to the overall risk [30].

The hospital-level covariates included in these models were: medical support (specialist versus general practitioner), years of operation (≥2 vs <2), nurses’ weekly working hours per hospital bed (>12 vs ≤12), and patients seen in a year (≥150 vs <150). The other attributes of CHs were constant or heavily skewed across providers, and thus excluded from multivariable analyses.

Due to concerns for over-fitting and misclassification, not all patient-level characteristics have been included in the models. A subset of all candidate variables, including demographics, source of admission, Barthel score on admission, hip fracture, and Elixhauser conditions, was preliminary chosen for inclusion in regression models using an automated selection method which is described in detail elsewhere [31, 32]. Scores on admission have been included in all of the Bayesian multilevel models for Barthel improvement, including the first, empty model.

All covariates were mean-centred to mitigate multi-collinearity and to allow for blocking of parameters in MCMC estimation. All analyses were carried out using the Stata 15 software (StataCorp. 2017. *Stata Statistical Software: Release 15*. College Station, TX: StataCorp LP).

### Ethics statement

In Italy, anonymous administrative data-gathering is subject to the law *Protection of individuals and other subjects with regard to the processing of personal data, ACT № 675 of 31.12.1996* (amended by Legislative Decree № 123 of 09.05.1997, № 255 of 28.07.1997, № 135 of 08.05.1998, № 171 of 13.05.1998, № 389 of 6.11.1998, № 51 of 26.02.1999, № 135 of 11.05.1999, № 281 of 30.07.1999, № 282 of 30.07.1999 and № 467 of 28.12.2001) (http://www.privacy.it/legge675encoord.html).

Data are pseudonymised on a regular basis at the regional statistical office, where each patient is assigned a unique identifier that eliminates the ability to trace the patient’s identity or other sensitive data. Pseudonymised administrative data can be used without a specific written informed consent when patient information is collected for health-care management and health-care quality evaluation and improvement (according to art. 110 on medical and biomedical and epidemiological research, Legislation Decree 101/2018).

Patients and the public were not involved in the design or planning of the study. All procedures performed in this study were in accordance with the 1964 Helsinki Declaration and its later amendments.

## RESULTS

### Patient case mix

A total of 3059 patients were discharged from the 16 CHs of Emilia-Romagna during the study period. Of these, 280 (9.2%) were aged <65 years and were excluded from the analyses. Because of missing data, we operated the additional exclusion of 292 (10.5%) patients referring to 3 non-respondent CHs. See supplementary Table S1 for a comparison of patient characteristics between respondent and non-respondent CHs: patients referring to non-respondent hospitals were more independent on admission (mean Barthel score: 42.0 vs. 30.3, P-value < 0.001); no other significant differences were observed.

The characteristics of the 2487 patients included in the study, overall and by CH, are summarized in Table 1: except patient demographics, the case mix appeared to be different across the 13 providers. The distribution of single clinical conditions identified with the Elixhauser method, overall and by CH, is presented in supplementary Table S2.

**Table 1.**
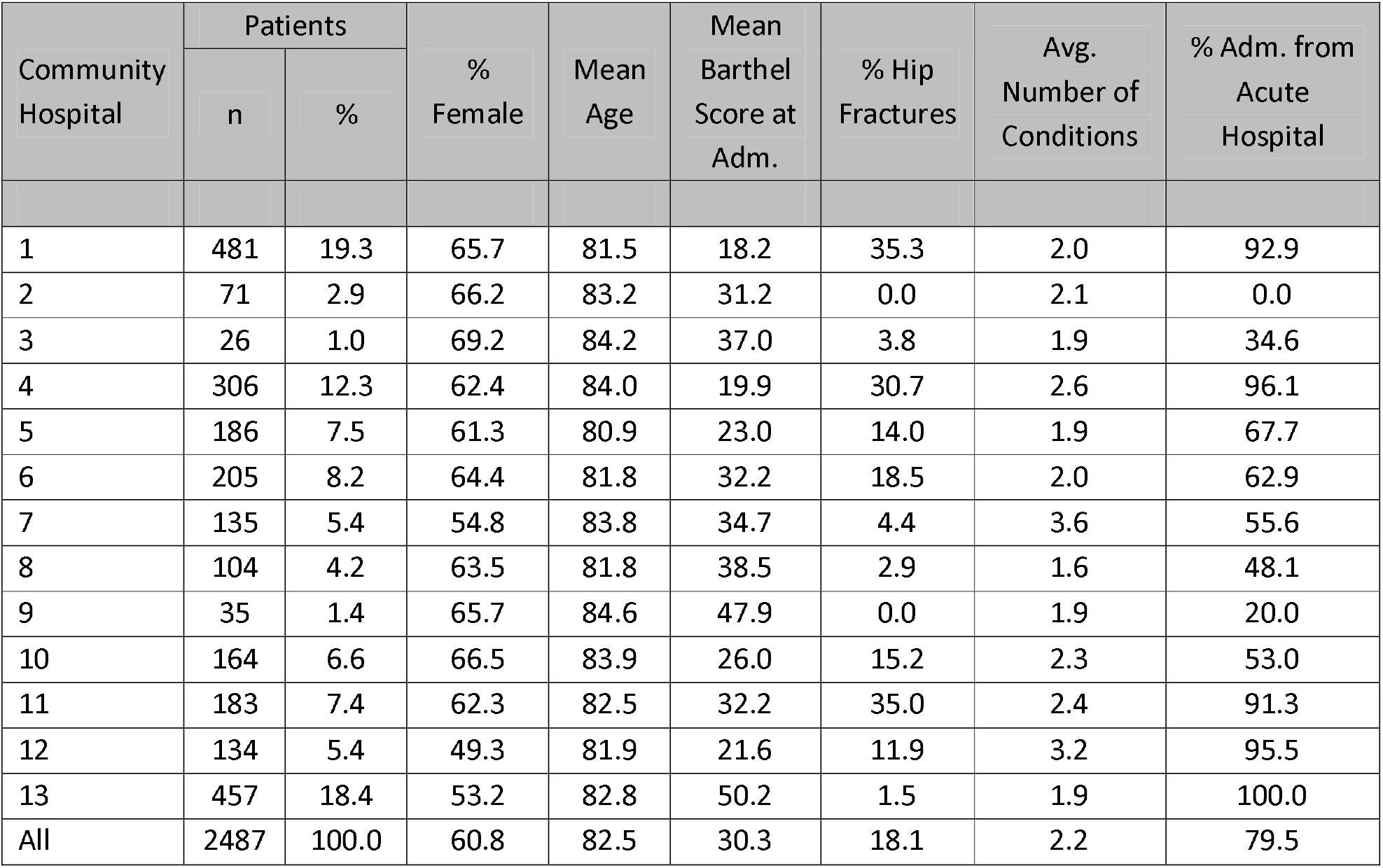
Characteristics of the 2487 study patients, overall and by community hospital, Emilia-Romagna, year 2017.

| Community Hospital | Patients |  | % Female | Mean Age | Mean Barthel Score at Adm. | % Hip Fractures | Avg. Number of Conditions | % Adm. from Acute Hospital |
| --- | --- | --- | --- | --- | --- | --- | --- | --- |
|  | n | % |  |  |  |  |  |  |
| 1 | 481 | 19.3 | 65.7 | 81.5 | 18.2 | 35.3 | 2.0 | 92.9 |
| 2 | 71 | 2.9 | 66.2 | 83.2 | 31.2 | 0.0 | 2.1 | 0.0 |
| 3 | 26 | 1.0 | 69.2 | 84.2 | 37.0 | 3.8 | 1.9 | 34.6 |
| 4 | 306 | 12.3 | 62.4 | 84.0 | 19.9 | 30.7 | 2.6 | 96.1 |
| 5 | 186 | 7.5 | 61.3 | 80.9 | 23.0 | 14.0 | 1.9 | 67.7 |
| 6 | 205 | 8.2 | 64.4 | 81.8 | 32.2 | 18.5 | 2.0 | 62.9 |
| 7 | 135 | 5.4 | 54.8 | 83.8 | 34.7 | 4.4 | 3.6 | 55.6 |
| 8 | 104 | 4.2 | 63.5 | 81.8 | 38.5 | 2.9 | 1.6 | 48.1 |
| 9 | 35 | 1.4 | 65.7 | 84.6 | 47.9 | 0.0 | 1.9 | 20.0 |
| 10 | 164 | 6.6 | 66.5 | 83.9 | 26.0 | 15.2 | 2.3 | 53.0 |
| 11 | 183 | 7.4 | 62.3 | 82.5 | 32.2 | 35.0 | 2.4 | 91.3 |
| 12 | 134 | 5.4 | 49.3 | 81.9 | 21.6 | 11.9 | 3.2 | 95.5 |
| 13 | 457 | 18.4 | 53.2 | 82.8 | 50.2 | 1.5 | 1.9 | 100.0 |
| All | 2487 | 100.0 | 60.8 | 82.5 | 30.3 | 18.1 | 2.2 | 79.5 |

### Study outcomes

After excluding scores on admission >90 (n = 116), we found 1261 patients with a change in Barthel index ≥10 (53.2%); after the exclusion of deaths during hospital stay (n = 158), we observed 262 transfers to acute care hospital (11.2%) and 1049 hospital stays ≥20 days (45.0%). The maximum length of stay in our data was 5 months. The observed hospital-specific outcome rates are presented in Table 2.

**Table 2.**
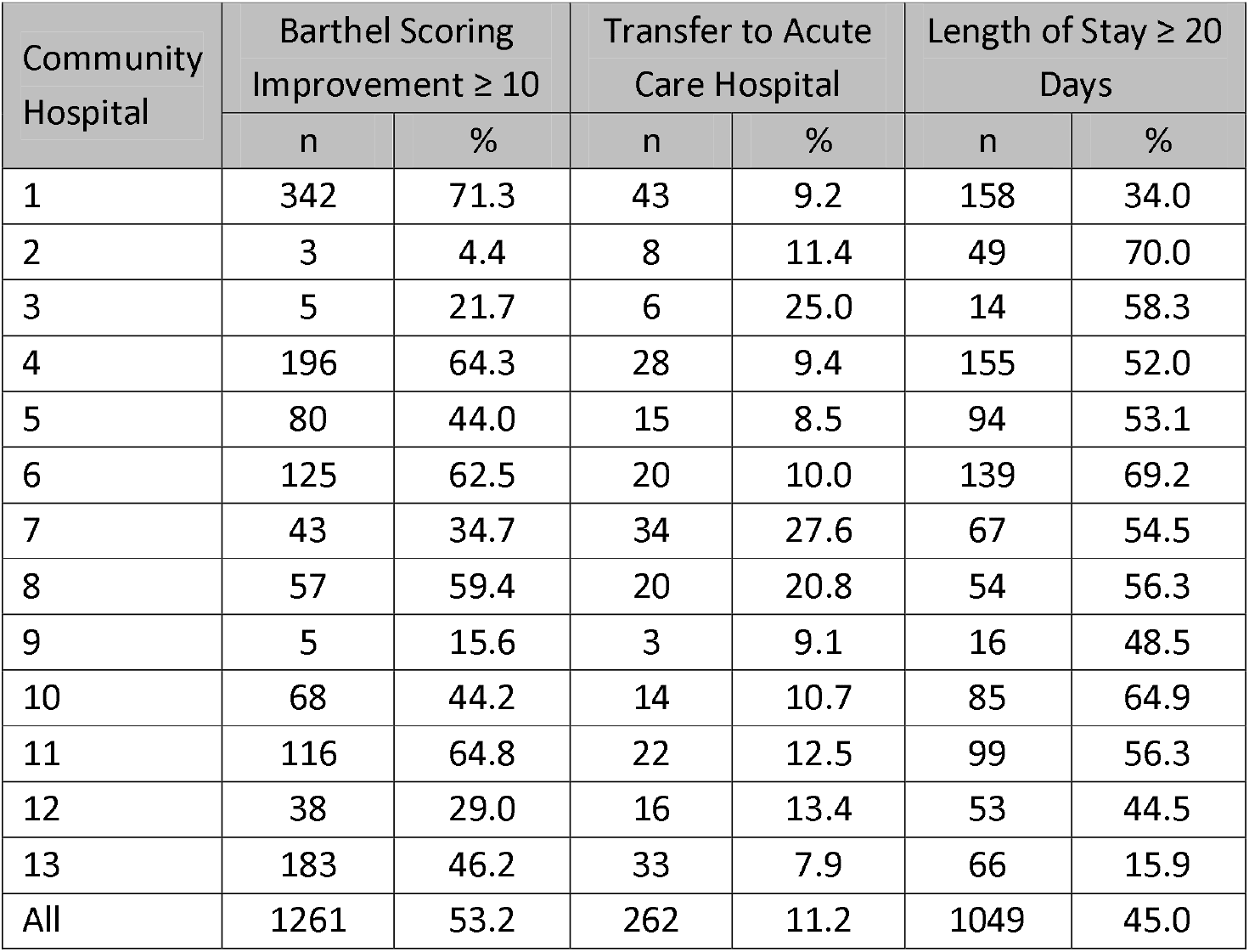
Rates of Barthel scoring improvement ≥10, transfer to acute care hospital, and length of stay ≥20 days in 13 community hospitals of Emilia-Romagna, year 2017.

| Community Hospital | Barthel Scoring Improvement $\geq 10$ | | Transfer to Acute Care Hospital | | Length of Stay $\geq 20$ Days | |
| --- | --- | --- | --- | --- | --- | --- |
|  | n | % | n | % | n | % |
| 1 | 342 | 71.3 | 43 | 9.2 | 158 | 34.0 |
| 2 | 3 | 4.4 | 8 | 11.4 | 49 | 70.0 |
| 3 | 5 | 21.7 | 6 | 25.0 | 14 | 58.3 |
| 4 | 196 | 64.3 | 28 | 9.4 | 155 | 52.0 |
| 5 | 80 | 44.0 | 15 | 8.5 | 94 | 53.1 |
| 6 | 125 | 62.5 | 20 | 10.0 | 139 | 69.2 |
| 7 | 43 | 34.7 | 34 | 27.6 | 67 | 54.5 |
| 8 | 57 | 59.4 | 20 | 20.8 | 54 | 56.3 |
| 9 | 5 | 15.6 | 3 | 9.1 | 16 | 48.5 |
| 10 | 68 | 44.2 | 14 | 10.7 | 85 | 64.9 |
| 11 | 116 | 64.8 | 22 | 12.5 | 99 | 56.3 |
| 12 | 38 | 29.0 | 16 | 13.4 | 53 | 44.5 |
| 13 | 183 | 46.2 | 33 | 7.9 | 66 | 15.9 |
| All | 1261 | 53.2 | 262 | 11.2 | 1049 | 45.0 |

### Skill mix and processes of care of community hospitals

Of the 13 CHs participating in this study (249 beds), 7 had been operating for ≥2 years and 7 admitted ≥150 patients over the study period. One CH was led by a teaching hospital, while the other 12 were led by local health-care authorities. Nine were located in health-care homes, 3 were located in acute hospitals and one was located in a residential facility. Eight CHs were led by GPs and 5 by specialists.

Clinical activity was organized differently across the CHs. Seven had GPs present in the facility on weekday mornings; in 5 of these, GPs were on call in the afternoon. In 5 CHs, care was ensured by specialists; in the smallest one, GPs offered medical consultation only through phone calls. In all CHs, specialists or first-aid physicians guaranteed clinical activity on weekends, off days and nights. Specialist medical advice was available in all CHs when needed.

Nurses’ weekly working hours per hospital bed were on average 11.4 hours (7 CHs ≤12 hours). Rehabilitation was delivered by physiotherapists in 12 CHs and in 11 a gym was present.

### Association of patient case mix with the study outcomes

Results from multilevel regression analyses are presented in Table 3. There is evidence that female sex, hip fracture, and admission from acute care hospital were associated with greater improvements in Barthel scores during hospital stay. On the contrary, the presence of specific clinical conditions (i.e., metastatic tumours, cerebrovascular diseases and dementia) and older age were associated with lower improvements.

**Table 3.** Results from Bayesian multilevel logistic regression analysis: association of patient case mix with the study outcomes, Emilia-Romagna, year 2017.

| Predictors | Barthel Scoring Improvement $\geq 10$ | | Transfer to Acute Care Hospital | | Length of Stay $\geq 20$ Days | |
| --- | --- | --- | --- | --- | --- | --- |
|  | Posterior OR | 95% Cred. Interval | Posterior OR | 95% Cred. Interval | Posterior OR | 95% Cred. Interval |
| Sex (female vs. male) | 1.22 | 1.001-1.47 | 0.83 | 0.63-1.09 | 1.30 | 1.07-1.55 |
| Age (1-year increase) | 0.95 | 0.94-0.96 | 0.99 | 0.97-1.003 | 0.99 | 0.98-1.01 |
| Barthel score at admission (1-point increase) | 1.02 | 1.01-1.02 | 0.99 | 0.98-0.995 | 0.99 | 0.986-0.993 |
| Squared-Barthel score at admission (1-point increase) | 0.999 | 0.999-0.999 | N/A |  | N/A |  |
| Hip fracture | 1.73 | 1.31-2.25 | 0.70 | 0.45-1.01 | 1.42 | 1.09-1.78 |
| Chronic kidney disease | N/A |  | 1.90 | 1.28-2.70 | N/A |  |
| Metastatic cancer | 0.31 | 0.16-0.54 | N/A |  | N/A |  |
| Cerebrovascular diseases | 0.69 | 0.55-0.87 | N/A |  | 1.45 | 1.16-1.80 |
| Dementia | 0.63 | 0.49-0.81 | N/A |  | N/A |  |
| Admission source (hospital vs. home/residential facility) | 1.45 | 1.08-1.92 | N/A |  | N/A |  |
| *Variance Partition Coefficient (VPC)* | 0.262 |  | 0.062 |  | 0.147 |  |

**Note:** Estimates are not available (N/A) if the patient-level predictor was discarded in preliminary automated regression analyses.
**Abbreviations:** OR, odds ratio, N/A, not available.

As indicated by the linear and quadratic terms of baseline Barthel index, highest and lowest scores had less chances of improvement, compared with patients with a medium level of independence on admission (see supplementary Figure S1). When accounting for these variables, the VPC decreased from 0.319 to 0.262, suggesting that a portion of variability across hospitals was explained by the patient case mix (both VPCs are controlled for patient admission scores). As also illustrated in Figure 1 (Panel a), the dispersion of hospital-specific random effects around the mean was lower after adjustment for patient characteristics. However, the presence of 6 outliers (i.e., providers whose 95% credible intervals do not include 1) suggests that hospital effects still accounted for significant variability in Barthel scoring improvement.

**Figure 1.**
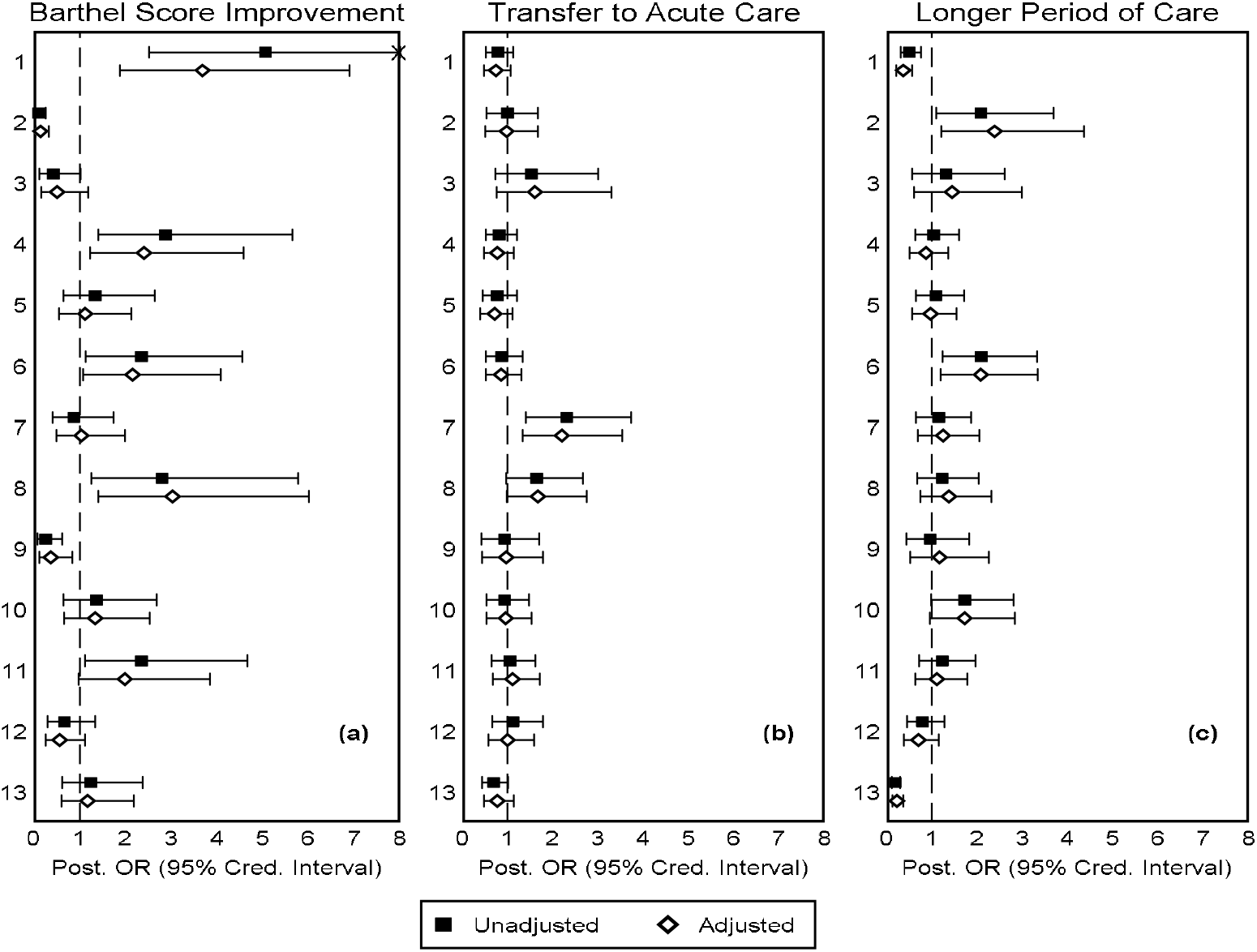
Results from Bayesian multilevel logistic regression models for (a) Barthel improvement ≥10 points, (b) transfer to acute care hospital, and (c) length of stay ≥20 days: hospital-specific odds ratios (ORs), unadjusted and adjusted for patient case mix, Emilia-Romagna, year 2017. Dashed line indicates the null value of 1 (no difference compared to the overall average). **Note:** Posterior ORs of Barthel scoring improvement were adjusted for sex, age, squared Barthel score on admission, hip fracture, metastatic cancer, cerebrovascular diseases, dementia, and admission source. Posterior ORs of transfer to acute care were adjusted for sex, age, Barthel score on admission, hip fracture, and chronic kidney disease. Posterior ORs of longer period of care were adjusted for sex, age, Barthel score on admission, hip fracture, and cerebrovascular diseases. The “unadjusted” posterior ORs of Barthel improvement were controlled for squared Barthel score on admission.

As shown in Table 3, less independent patients (as indicated by lower Barthel scores on admission) and patients with chronic kidney disease were associated with higher risk of transfers to acute care settings. Less independent patients, hip fractures and cerebrovascular conditions were also associated with longer periods of intermediate care. However, the informative contribution of these covariates to the variability across providers was of no account (hospital transfer-VPC 0.059 to 0.062; longer stay-VPC 0.138 to 0.147). It should be noted too that the variability across providers in these two outcomes was lower than the variability in Barthel improvements (Figure 1, Panels b, c).

### Association skill mix and process of care with the study outcomes

None of the parameters involving skill mix and processes of care were significantly associated with the study outcomes (Table 4). Of note, the VPCs resulting from adding these variables to the models were virtually coincident with those from previous analyses (Barthel improvement-VPC: 0.262 to 0.279; hospital transfer-VPC: 0.062 to 0.055; longer stay-VPC: 0.147 to 0.143).

**Table 4.** Results from Bayesian multilevel logistic regression analysis: association of skill mix and care processes of community hospitals with the study outcomes, Emilia-Romagna, year 2017.

| Predictors | Barthel Scoring Improvement $\geq 10$ | | Transfer to Acute Care Hospital | | Length of Stay $\geq 20$ Days | |
| --- | --- | --- | --- | --- | --- | --- |
|  | Posterior OR* | 95% Cred. Interval | Posterior OR* | 95% Cred. Interval | Posterior OR* | 95% Cred. Interval |
| Medical support (specialist vs GP) | 1.76 | 0.28-8.46 | 0.79 | 0.45-1.31 | 0.51 | 0.15-1.22 |
| Years of operation ($\geq 2$ vs $< 2$) | 1.12 | 0.23-3.25 | 0.95 | 0.55-1.51 | 1.24 | 0.46-2.85 |
| Nurses' working hours/week/bed ($> 12$ vs $\leq 12$) | 0.89 | 0.18-2.77 | 1.20 | 0.70-2.00 | 1.39 | 0.47-3.26 |
| Patients/year ($\geq 150$ vs $< 150$) | 4.21 | 0.47-15.70 | 0.61 | 0.33-1.07 | 1.02 | 0.31-2.54 |
| *Variance Partition Coefficient (VPC)* | 0.279 |  | 0.055 |  | 0.143 |  |

\* OR estimates are adjusted for patient-level characteristics listed in Table 3.
**Abbreviations:** OR, odds ratio, N/A, not available.

## DISCUSSION

In this population-based observational study, we found that female sex, younger age, hip fracture and admission from acute care hospital were associated with greater improvement in Barthel scores; on the contrary, metastatic tumours, cerebrovascular diseases and dementia were associated with lower improvement. Females, hip fractures and cerebrovascular diseases were associated with longer hospital stays. As regards the impact of skill mix and care processes on the study outcomes, no statistically significant association was found.

The Barthel index is usually adopted as outcome measure for intermediate care facilities: the UK National Audit of Intermediate Care uses it to evaluate patients’ improvements between CH admission and discharge. In our analysis, 53.2% of the patients had an improvement in their functional ability during hospitalization (≥10). According to Dixon et al. [23], less independent patients on admission (i.e., patients with a lower Barthel score) were associated with greater improvement in Barthel scoring; our evidence showed that highest and lowest Barthel scores had less chances of improvement compared with patients that presented with a medium level of independence.

Furthermore, in our study 88.8% of the patients were discharged at home to continue treatment closer to family or in a familiar environment and only 45% had a hospital stay ≥20 days. These data suggest that CHs could be the appropriate setting of care for frail and older patients needing rehabilitation or low intensity care, with the purpose of enabling them to experience a better quality of life after discharge from an acute hospital or to prevent them from being hospitalized when not necessary. Other studies too have shown that intermediate care can enable patients to regain abilities in daily living, decrease readmissions and reduce mortality [33 - 37]. Overall, the case mix was different across the 13 CHs, but the study patients showed similar demographic characteristics and most of them were frail, elderly, with multiple conditions and with significant functional decline in the activities of daily living. As other authors have pointed out, resource consumption and poor outcomes are significantly higher for these patients due to their clinical and socio-demographic conditions [38, 39]. Our findings too suggest that clinical outcomes are negatively affected by age and multimorbidity, which is more frequent with increasing age [40]. A possible explanation for the case mix heterogeneity among CHs is that each service needs to be adapted to the catchment area in which it is implemented, responding to the different needs of the population it serves.

The CHs’ organizational model, similarly to other intermediate care services, faced the challenge of adapting to the local context and responding to the different needs of the populations by drawing patient centred services, involving an adequate skill mix and building an integrated system among different settings, disciplines and professions [41 - 42]. To do that, different organizational models were implemented across the CHs of Emilia-Romagna: the heterogeneity of case mix served and skill mix required represents both a strength (i.e., capacity of adapting to the local context) and a weakness (i.e., difficulty of standardizing a model that can work everywhere) of these services. In Emilia-Romagna, we reported that clinical responsibilities in the CHs more frequently involved physicians from various disciplines, contrary to what happens in other countries where the involvement of doctors is less diverse [43 - 48]. In 8 out of 13 CHs, GPs had clinical responsibilities; specialists were available for clinical consultation in all the structures, and in 5 of them they also had clinical accountability and were present in the structure at all times. Nurses held great managerial and patient-related tasks, similarly to other countries. [44, 49]. All professionals involved worked together as a multidisciplinary/multiprofessional team. This could be seen as an advantage per se, because collaborative capacity is fundamental for integrated care services. However, developing effective teams and organizations requires and the personnel needs to build relationships between partners in care to establish trust and willingness to take shared accountability for outcomes. Accordingly, the growth of competencies to deliver integrated care should start with accurate development and training of health professionals [24].

Our analysis showed no statistically significant association of CH skill mix and care process with the study outcomes. However, our results indicate that in 6 CHs a part of variability in the Barthel scoring improvements could be attributed to specific characteristics of the services that we were not able to detect with our survey. New research on intermediate care, using mixed quantitative-qualitative methods such as the positive deviant approach [50], could help highlight specific aspects of skill mix and processes of care that impact favourably on the patient’s level of independence.

We argue that another important condition that should be investigated as both access condition and outcome is frailty. The continuous rising of frailty makes it harder to meet the health and social needs of the elderly [51], and an association with adverse outcome is reported in various studies [52 - 56]. For this reason, knowing which patient domains (i.e., physical, cognitive, social and psychological) mostly drive the path to loss of function would serve as a proxy for health-care utilization and improve the quality of patient-centred care [57].

A strength of this study was the possibility to link subject-level data using the unique patient identifier. We also created an ad hoc questionnaire with the purpose of investigating CHs’ organizational models, giving us the opportunity to analyse specific features such as nurses’ and support staff’s weekly working hours and specific medical support. Limitations to the study were the small number of CHs investigated, although the response rate was high. Because we did not follow up patients after discharge, we cannot conclude that favourable study outcomes were associated with a lower risk of hospital readmission or death. Lastly, the Barthel index is a convenient measurement scale to assess physical functional disability, but fails to capture changes in functional ability related to general health, communication status and psychological status.

## CONCLUSIONS

CHs could be the right setting of care for elderly frail patients with complex health and social needs. The diverse organization of CHs makes it difficult to summarize its effectiveness, as also reported by other authors. Furthermore, the methods and outcome measures currently used might not be appropriate to weigh the relative contribution of case mix, skill mix and care processes to the patient’s health and wellbeing. Novel approaches, such as mixed quantitative-qualitative methods, could be used to evaluate thoroughly the outcomes of intermediate care services. Moreover, increasing awareness of frailty among providers and fostering teamwork could make care more tailored to the needs of patients.

## Data Availability

Data sources were the regional Community Hospitals informative system (SIRCO), hospital discharge records and an ad hoc questionnaire for Community Hospitals

## DECLARATIONS

### Availability of data and material

Data can be available if requested.

### Ethics approval

Not applicable.

## SUPPLEMENTARY TABLES

**Table S1.**
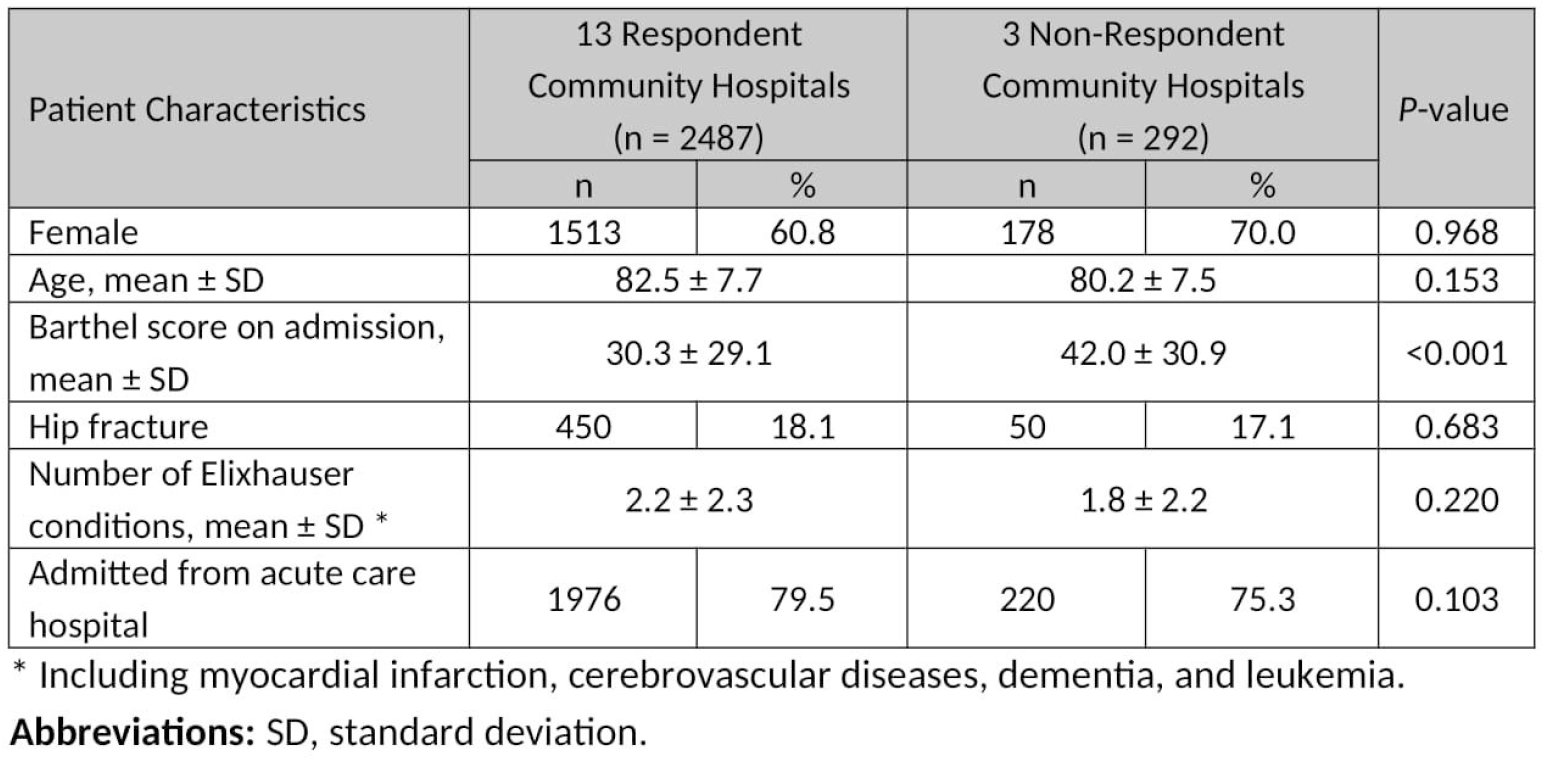
Population characteristics in respondent and non-respondent community hospitals of Emilia-Romagna, year 2017.

| Patient Characteristics | 13 Respondent Community Hospitals (n = 2487) |  | 3 Non-Respondent Community Hospitals (n = 292) |  | P-value |
| --- | --- | --- | --- | --- | --- |
|  | n | % | n | % |  |
| Female | 1513 | 60.8 | 178 | 70.0 | 0.968 |
| Age, mean ± SD | 82.5 ± 7.7 | | 80.2 ± 7.5 | | 0.153 |
| Barthel score on admission, mean ± SD | 30.3 ± 29.1 | | 42.0 ± 30.9 | | <0.001 |
| Hip fracture | 450 | 18.1 | 50 | 17.1 | 0.683 |
| Number of Elixhauser conditions, mean ± SD * | 2.2 ± 2.3 | | 1.8 ± 2.2 | | 0.220 |
| Admitted from acute care hospital | 1976 | 79.5 | 220 | 75.3 | 0.103 |

\* Including myocardial infarction, cerebrovascular diseases, dementia, and leukemia.
**Abbreviations:** SD, standard deviation.

**Table S2.**
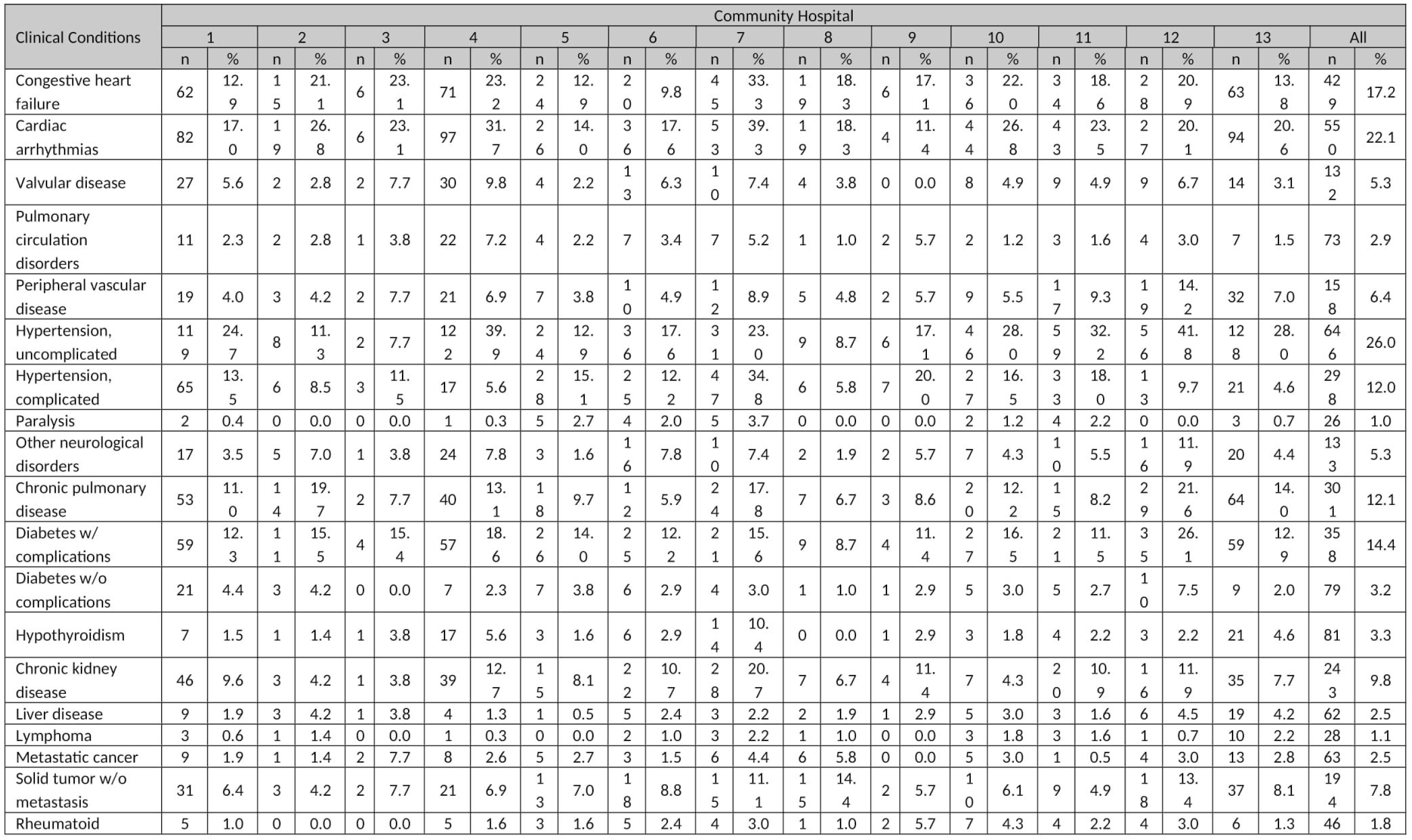
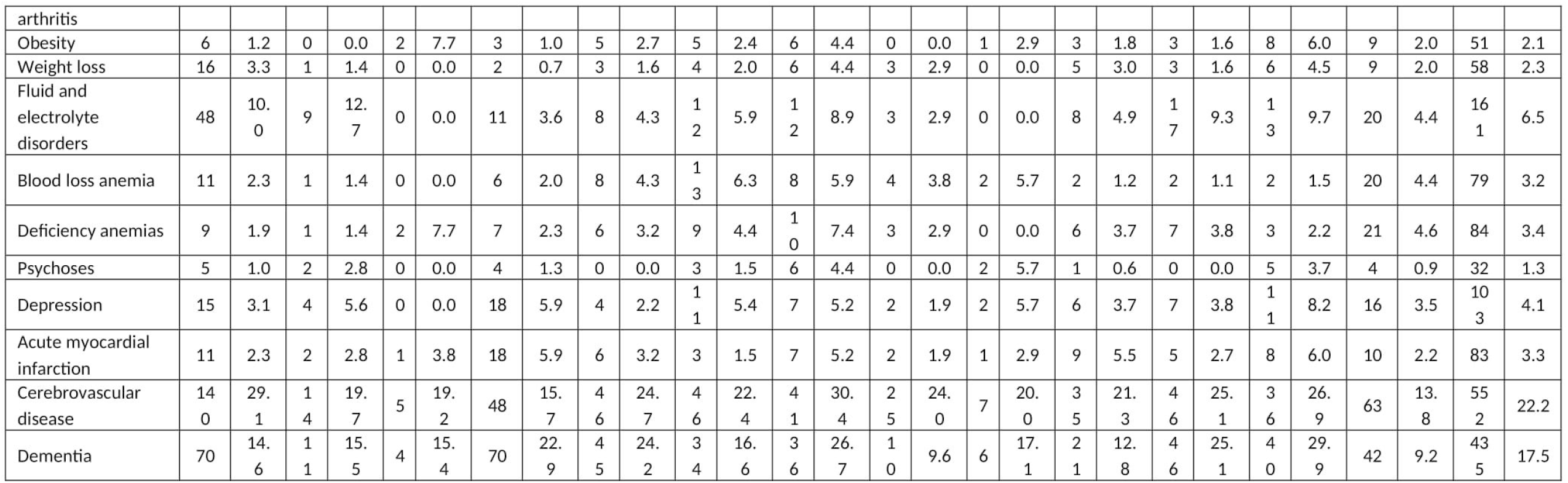
Distribution of clinical conditions occurring 3-years prior to the community hospital admission, overall and by community hospital of Emilia-Romagna, year 2017. Conditions with prevalence <1%are not reported.

**Figure S1.**
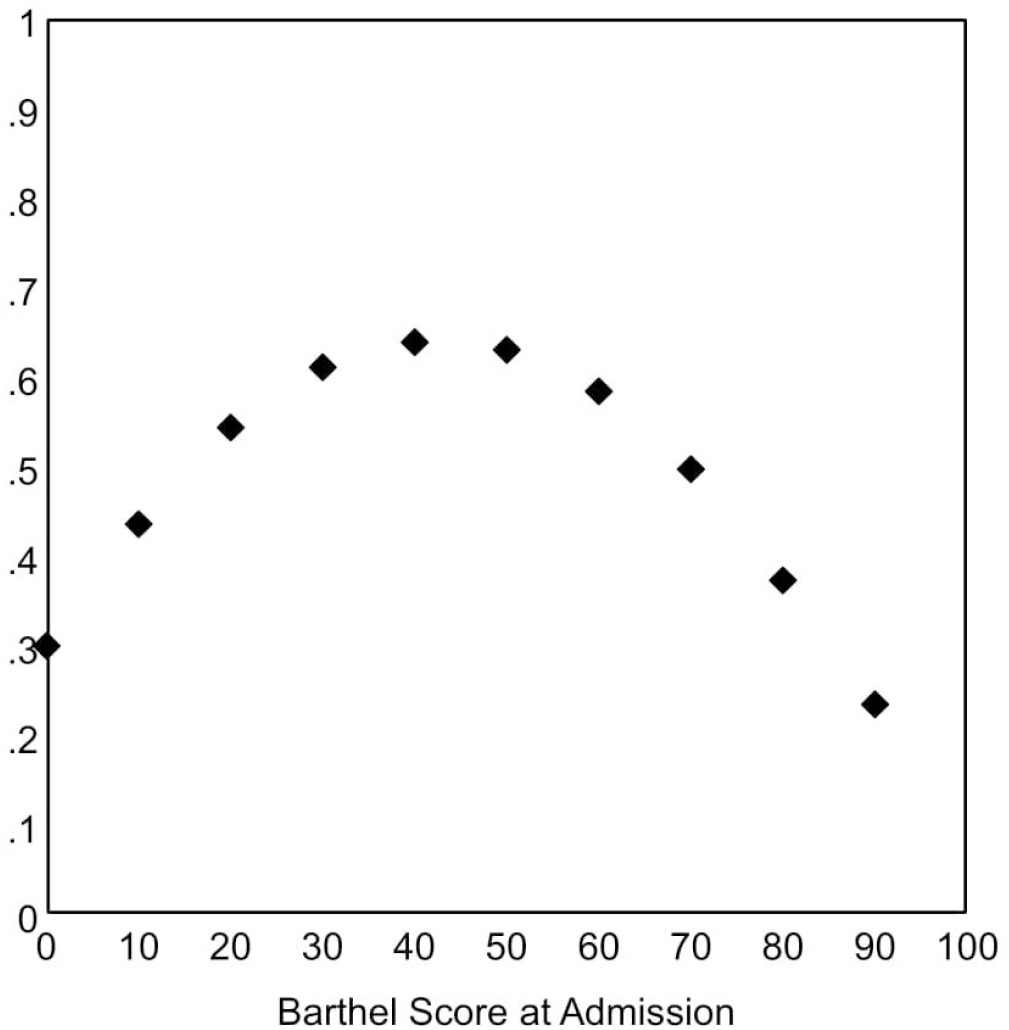
Results from Bayesian multilevel logistic regression models for Barthel improvement ≥10 points: predicted probability of improvement as a function of Barthel score on admission. Emilia-Romagna, year 2017. **Note:** Predicted probabilities were adjusted for sex, age, hip fracture, metastatic cancer, cerebrovascular diseases, dementia, and admission source.

